# Polysubstance addiction and psychiatric, somatic comorbidities among 7,989 individuals with cocaine use disorder: a latent class analysis

**DOI:** 10.1101/2023.02.08.23285653

**Authors:** Brendan Stiltner, Robert H. Pietrzak, Daniel S. Tylee, Yaira Z. Nunez, Keyrun Adhikari, Henry R. Kranzler, Joel Gelernter, Renato Polimanti

## Abstract

**Aims:** We performed a latent class analysis (LCA) in a sample ascertained for addiction phenotypes to investigate cocaine use disorder (CoUD) subgroups related to polysubstance addiction (PSA) patterns and characterized their differences with respect to psychiatric and somatic comorbidities.

**Design:** Cross-sectional study

**Setting:** United States

**Participants:** Adult participants aged 18-76, 39% female, 47% African American, 36% European American with a lifetime DSM-5 diagnosis of CoUD (N=7,989) enrolled in the Yale-Penn cohort. The control group included 2,952 Yale-Penn participants who did not meet for alcohol, cannabis, cocaine, opioid, or tobacco use disorders.

**Measurements:** Psychiatric disorders and related traits were assessed via the Semi-structured Assessment for Drug Dependence and Alcoholism. These features included substance use disorders (SUD), family history of substance use, sociodemographic information, traumatic events, suicidal behaviors, psychopathology, and medical history. LCA was conducted using diagnoses and diagnostic criteria of alcohol, cannabis, opioid, and tobacco use disorders.

**Findings:** Our LCA identified three subgroups of PSA (i.e., low, 17%; intermediate, 38%; high, 45%) among 7,989 CoUD participants. While these subgroups varied by age, sex, and racial-ethnic distribution (p<0.001), there was no difference on education or income (p>0.05). After accounting for sex, age, and race-ethnicity, the CoUD subgroup with high PSA had higher odds of antisocial personality disorder (OR=21.96 vs. 6.39, difference-p=8.08×10^−6^), agoraphobia (OR=4.58 vs. 2.05, difference-p=7.04×10^−4^), mixed bipolar episode (OR=10.36 vs. 2.61, difference-p=7.04×10^−4^), posttraumatic stress disorder (OR=11.54 vs. 5.86, difference-p=2.67×10^−4^), antidepressant medication use (OR=13.49 vs. 8.02, difference-p=1.42×10^−4^), and sexually transmitted diseases (OR=5.92 vs. 3.38, difference-p=1.81×10^−5^) than the low-PSA CoUD subgroup.

**Conclusions:** We found different patterns of PSA in association with psychiatric and somatic comorbidities among CoUD cases within the Yale-Penn cohort. These findings underscore the importance of modeling PSA severity and comorbidities when examining the clinical, molecular, and neuroimaging correlates of CoUD.

## INTRODUCTION

Substance use disorders (SUDs) represent heterogeneous patterns of behavioral, cognitive, and physiological features associated with the chronic use of a substance, despite problems that arise from its use (1). Among SUDs, cocaine use disorder (CoUD) contributes to significant morbidity, mortality, personal, and healthcare expense, posing a considerable public health problem (2). In 2019, nearly 5.5 million people in the United States reported using cocaine in the past year and 1 million met criteria for CoUD (3).

Polysubstance use and addiction are defined as the consumption and abuse of more than one substance over a short period of time or concurrently (4). These are both common and associated with adverse consequences (5). People who use and abuse multiple substances have worse treatment outcomes (6, 7), higher risk of mortality compared to people that use one substance (8), and are statistically more likely to experience an overdose, violence, and accidental injuries than those that use one substance (9). With respect to the risk of polysubstance use among cocaine users, 72.7% of cocaine-involved deaths registered in 2017 involved opioids (10).

Modeling patterns of polysubstance addiction (PSA) among substance users can help to define high-risk groups within diagnostic boundaries. Latent class analysis (LCA) is a statistical method that can be used to identify groups of people (referred to as classes) based on similar patterns of manifest response variables (11). This allows one to identify more homogenous groups within a heterogenous population (12). Studies have used LCA within samples of people who use substances or diagnosed with a SUD to evaluate patterns of association with comorbid SUDs and clinically relevant variables (13, 14). Previous studies examined nationally Mrepresentative samples (15, 16) and treatment-seeking populations (17-19). However, these types of study designs feature limited sample sizes for rare SUDs such as CoUD.

Latent variable modeling approaches have also been used to identify classes of people with certain patterns of symptoms. The approach has been used in posttraumatic stress disorder (20, 21) and individuals reporting manic episodes (22). These studies revealed that different latent symptom typologies are differentially related to meaningful clinical and neurobiological markers. However, in SUDs, these approaches have focused mainly on alcohol (23, 24) and cannabis use disorders (25-27). To date, there have been few efforts to model the criteria of other SUDs (28, 29) and, to our knowledge, none have assessed PSA patterns in individuals with CoUD. However, previous research has identified latent classes of cocaine dependence based on other approaches (30, 31).

Using data from the large Yale-Penn cohort, we investigated PSA latent classes in 7,989 participants with lifetime DSM-5 CoUD diagnoses. Within these latent classes of CoUD, we also characterized symptom profiles and tested their associations with clinically meaningful mental and physical health variables. Our results show that clusters of CoUD-affected individuals have different patterns of PSA and of psychiatric and somatic comorbidities. These findings have important implications for modeling CoUD heterogeneity in studies of addiction-ascertained cohorts (e.g., molecular, brain imaging, and treatment investigations).

## METHODS AND MATERIALS

### Participants

Participants in this study are from the Yale-Penn cohort, which was recruited to investigate the genetics of substance use disorders and their comorbid conditions (32-37). Participants were recruited from five US sites: Yale School of Medicine (APT Foundation, New Haven, CT, USA), University of Connecticut Health Center (Farmington, CT, USA), University of Pennsylvania Perelman School of Medicine (Philadelphia, PA, USA), Medical University of South Carolina (Charleston, SC, USA), and McLean Hospital (Belmont, MA, USA). The studies were approved by the institutional review boards at each site and written informed consent was obtained from each participant.

### Measures

Participants were interviewed by trained personnel using the Semi-Structured Assessment for Drug Dependence and Alcoholism (SSADDA). The SSADDA produces lifetime diagnoses of substance use disorders and other mental illnesses (38, 39). The SSADDA includes items to diagnose substance dependence and abuse for the major substances of abuse, except for tobacco, for which there is no abuse diagnosis. In the present study, we derived lifetime DSM-5 diagnoses of SUD for alcohol, cannabis, cocaine, opioid, and tobacco. However, for DSM-5 tobacco use disorder, there are only 9 criteria available in the SSADDA. Additionally, we extracted from the SSADDA information regarding several psychiatric and behavioral traits. These included phenotypes relate to suicidality, family history of substance use, traumatic experiences, and psychopathology. Data related to socioeconomic status and physical health were also included in the analyses. Supplemental Table 1 describes the items included and how they were assessed.

### Data analyses

LCA was first performed including participants with a lifetime CoUD diagnosis (n=7,989) and using lifetime alcohol use disorder (AUD), cannabis use disorder (CaUD), opioid use disorder (OUD), and tobacco use disorder (TUD) as manifest variables. We assigned participants to the resulting classes if they had posterior probability ≥0.7 as previously proposed (40, 41). Next, within each diagnosis-based latent class, we performed LCAs, in which we analyzed the specific criteria for each SUD separately (alcohol, opioid, cannabis, and tobacco). To determine the best-fitting solution, we fit models beginning with 2 classes and increased the number of classes by one at each step. We selected the best-fitting solution based on model-fit statistics, size of the smallest class, and the interpretability and applicability of the model (42). Goodness-of-fit statistics included the Bayesian information criterion (BIC) and Akaike information criterion (AIC) (42). For the criteria-based classes, we selected models that produced classes with ≥ 5% of the sample of the diagnosis-based class. When there was disagreement between these indicators, we selected models with the highest entropy (43) and most significant bootstrap likelihood ratio test (BLRT) (42).

Next, we investigated differences between each diagnosis-based latent class and Yale-Penn participants who did not meet criteria for any of the 5 SUDs of interest (n=2,952). This analysis was performed using logistic regression models with age, sex, and self-reported race-ethnicity categories as covariates. The dependent variables included traits related to psychopathology, suicidality, traumatic experiences, family history of substance use, medical conditions, and socioeconomic status (SES). For variables on which multiple classes differed significantly from controls, we used z-tests to assess differences between the effect size of the associations observed with respect to the classes. All analyses were done in RStudio (44, 45). The poLCA package (46, 47) was used to conduct the LCAs and glca package (48) was used to test for significant differences between LCA models.

## RESULTS

### Sample Characteristics

Among 7,989 Yale-Penn participants with a lifetime diagnosis of CoUD, 39% were female, and the average age was 41 years (ranging from 18 to 76 years old, standard deviation = 9.6 years). The sample included mostly African Americans (47%) and European Americans (36%). With respect to comorbid SUDs, 82% met criteria for AUD, 57% for CaUD, 49%, for OUD, and 86% for TUD. With regard to family history of substance use, 51% reported being aware of at least one family member using any drugs or alcohol before the participant was 13 years of age. Participants with CoUD had family members who used cocaine (9%), heroin (5%), abused prescription drugs (4%), or used other illegal drugs (16%), and 47% and 65% were aware of family members who drank alcohol or smoked cigarettes, respectively. Table 1 summarizes the sample characteristics.

**Table 1.**
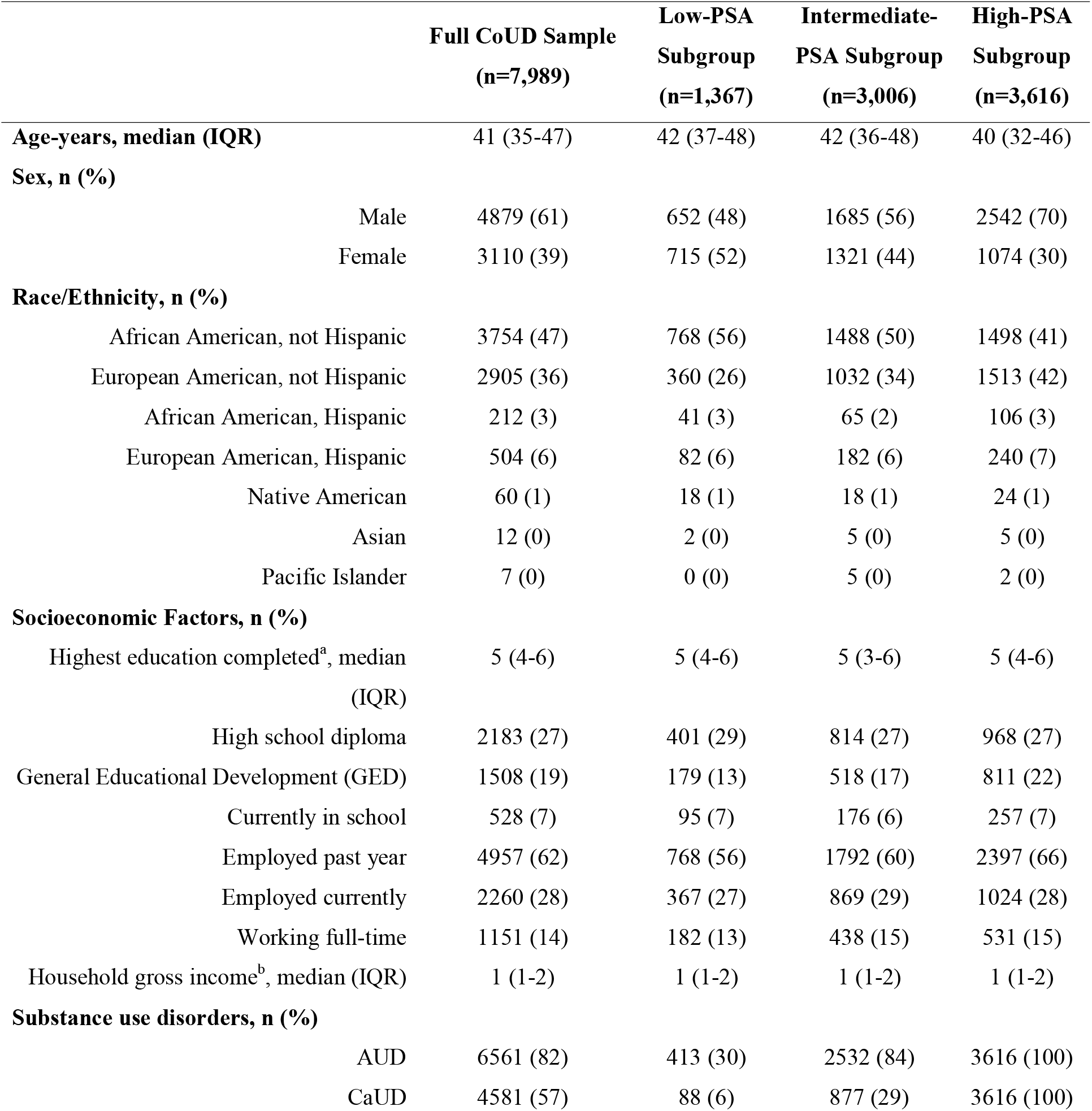
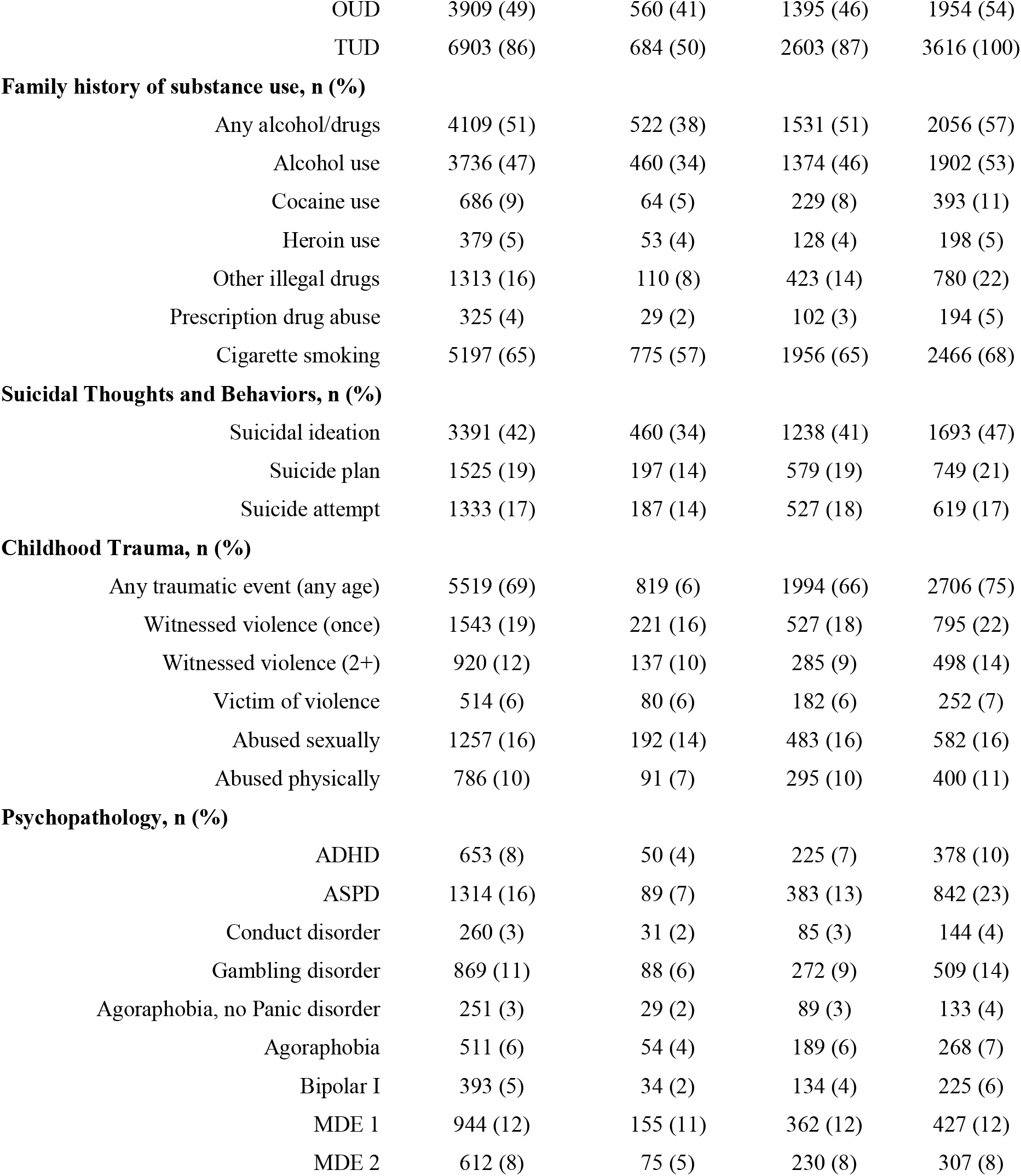
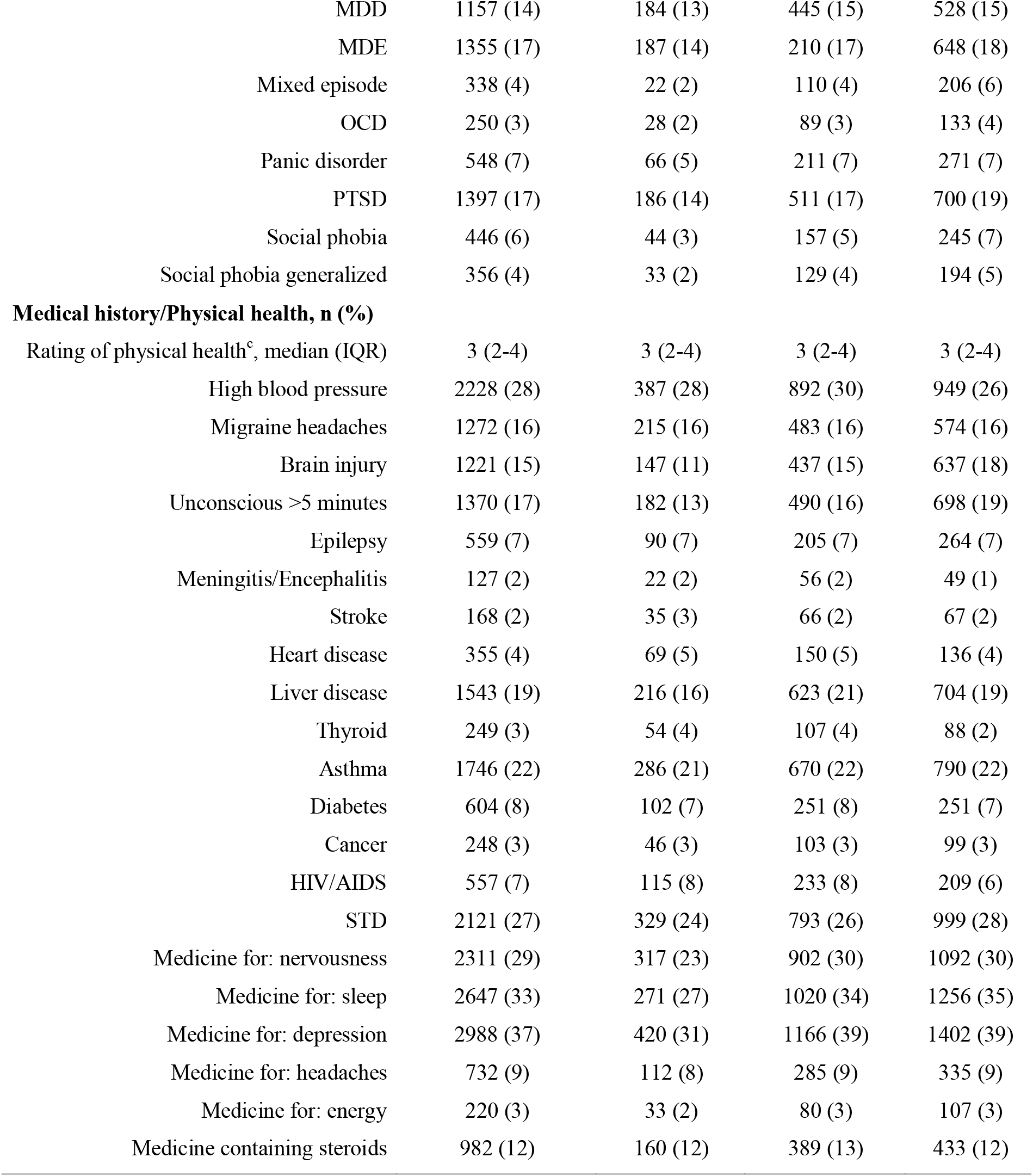
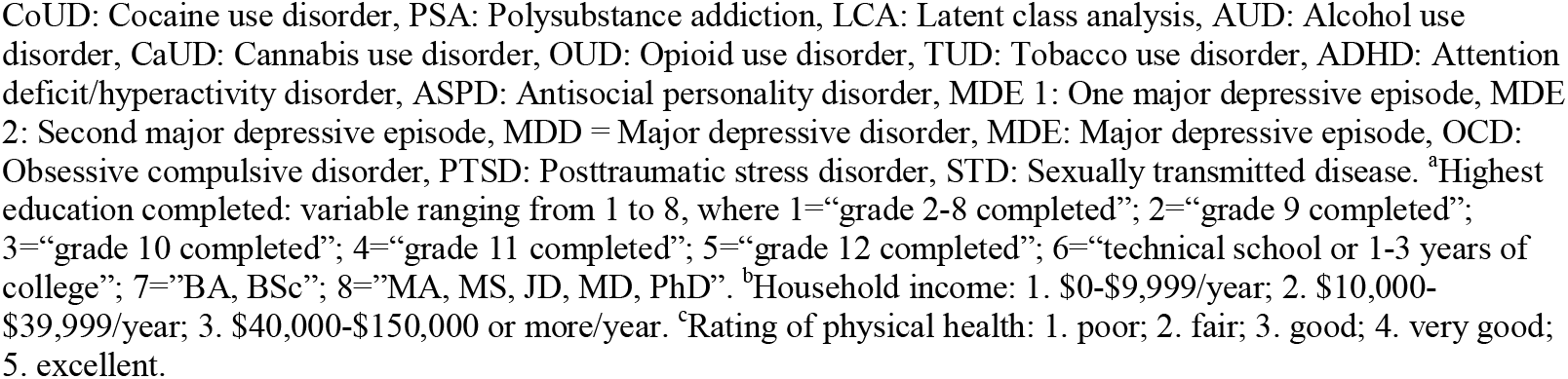
Characteristics of the full CoUD sample and diagnosis-based LCA subgroups based on AUD, CaUD, OUD, and TUD diagnoses.

### Diagnosis-Based Latent Class Analysis

Our initial LCA of CoUD-affected individuals was based on diagnoses of AUD, CaUD, OUD, and TUD. With only four dichotomous variables, we could identify only a two-class model (49). Considering a posterior probability ≥ 0.70, we assigned 1,367 individuals (17%) to Class-1 and 3,616 individuals (45%) to Class-2. No participant had a posterior probability greater than 0.7 for both classes. Conversely, 38% of the CoUD sample (n=3,006) had a posterior probability < 0.7 with respect to both diagnosis-based latent classes. Based on these results and the prevalence of other SUDs in these three groups (Table 1), we defined three CoUD subgroups: a low-PSA subgroup (i.e., diagnosis-based Class-1), an intermediate PSA subgroup (i.e., the group of participants with posterior probabilities < 0.7 for both diagnosis-based classes), and a high-PSA subgroup (i.e., diagnosis-based Class-2). Relative to the overall CoUD sample, the low-PSA subgroup was older (42.5 vs. 40.7 years of age, p<0.0001), had a higher proportion of females (52% vs. 39%, p<0.0001) and non-Hispanic African Americans (56% vs. 47%, p<0.0001), and a smaller proportion of fewer non-Hispanic European Americans (26% vs. 36%, p<0.001). An opposite pattern was present for the high-PSA subgroup, which was slightly younger (39.2 vs. 40.7 years of age, p<0.0001), had a smaller proportion of females (30% vs. 39%, p<0.0001) and non-Hispanic African Americans (41% vs. 47%, p<0.0001), and a larger proportion of non-Hispanic European Americans (42% vs. 36%, p<0.0001) than the overall CoUD sample. The intermediate-PSA subgroup showed characteristics intermediate between the two other subgroups and more similar to the overall CoUD sample. No differences were observed between the CoUD subgroups and the overall CoUD sample on education or income (p>0.05).

The distribution of SUD diagnoses for the low-PSA subgroup had lower prevalence of SUD diagnoses than the overall CoUD sample: 30% vs. 82% for AUD (p<0.0001); 6% vs. 57% for CaUD (p<0.0001); 41% vs. 49% for OUD (p<0.0001), and 50% vs. 80% for TUD (p<0.0001). Conversely, the high-PSA subgroup included higher rates SUD of comorbidity, with 100% of individuals meeting criteria for AUD (p<0.0001), CaUD (p<0.0001), and TUD (p<0.0001). The intermediate-PSA subgroup showed a similar distribution of AUD (84% vs. 82%, p<0.05), OUD (46% vs. 49%, p<0.05), and TUD (87% vs. 86%, p>0.05), but a much lower proportion of CaUD cases (29% vs. 57%, p<0.0001) than the overall CoUD sample. Supplemental Table 2 reports the SUD distribution in African Americans and European Americans across the overall CoUD sample and three CoUD subgroups.

### Criteria-Based Latent Class Analysis

To further investigate PSA patterns within each of the CoUD subgroups (Table 1), we conducted additional LCAs using DSM-5 diagnostic criteria for AUD, CaUD, OUD, and TUD. The number of diagnostic-criteria latent classes in each of the SUDs considered was defined based on AIC, BIC, BLRT, and entropy statistics (Supplemental Tables 3 to 5). Figure 1 summarizes the number of diagnostic-criteria latent classes, their posterior probabilities, and predicted class membership across the three CoUD subgroups. Considering the number of diagnostic-criteria latent classes, the low-PSA subgroup showed the best fit to the data with two diagnostic-criteria latent classes for CaUD, three for AUD and OUD, and four for TUD. Conversely, four or five diagnostic-criteria latent classes across the SUDs considered were the best fits for the intermediate and high-PSA subgroups.

**Figure 1.**
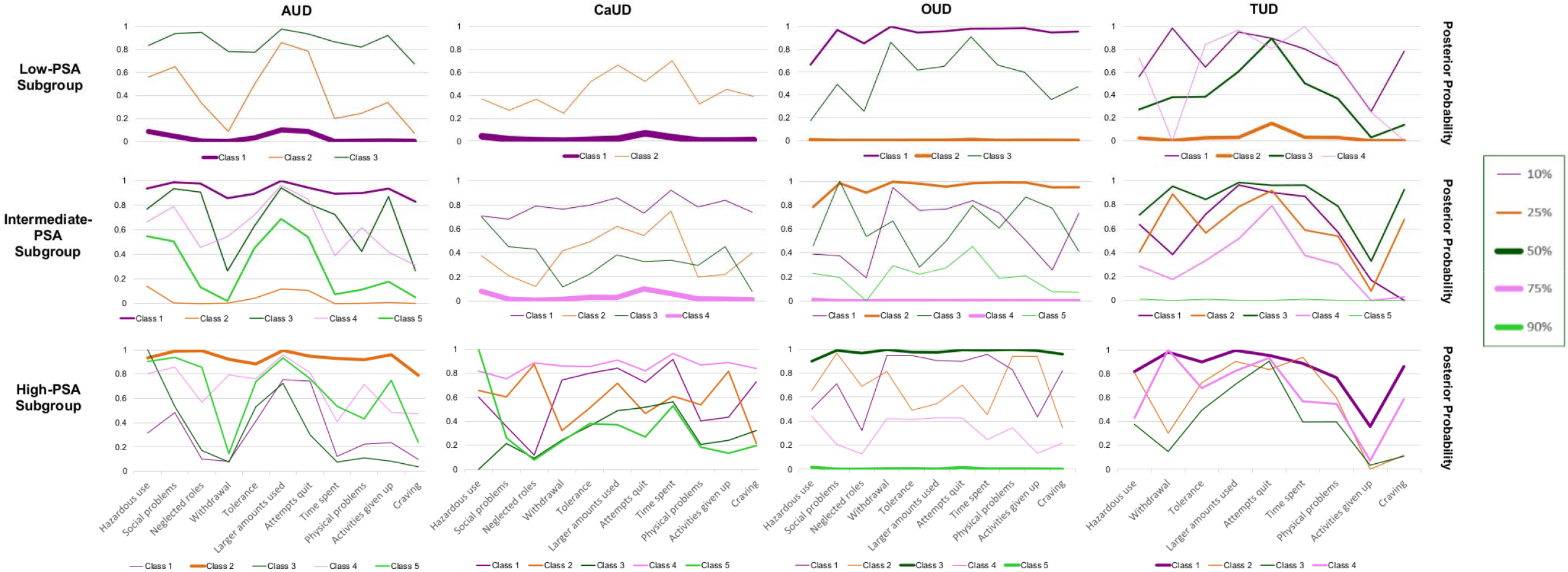
Patterns of the posterior probabilities of the criteria-based latent classes across CoUD subgroups. The posterior probabilities of the best-fit latent class models identified for AUD, CaUD, OUD, and TUD criteria with respect to the three CoUD subgroups. Line thickness corresponds to the proportion of the diagnosis-based subgroup that was stratified to a particular criteria-based class.

Considering the posterior probabilities and the predicted class memberships of the diagnostic-criteria latent classes, we observed unique PSA patterns across the CoUD subgroups. Specifically, most individuals included in the low-PSA subgroup were assigned to latent classes with <20% posterior probability of diagnostic criteria for AUD (predicted class membership=73%), CaUD (predicted class membership=95%), and TUD (predicted class membership=50%). Conversely, the high PSA subgroup showed 44% predicted class membership for a latent class with >78% posterior probability across AUD diagnostic criteria and 43% predicted class membership for a latent class with >76% posterior probability across TUD diagnostic criteria with the exception of the TUD criterion “Important activities are reduced or given up because of use.” With respect to CaUD, the high-PSA subgroup showed diagnostic-criteria latent classes (predicted class membership ranging from 16% to 25%) with different patterns of posterior probabilities. Participants in the intermediate-PSA subgroup were evenly distributed across diagnostic-criteria latent classes with different patterns of posterior probabilities for AUD and TUD. Conversely, 75% of this subgroup was predicted to be assigned to a latent class with <10% posterior probability of CaUD diagnostic criteria. Interestingly, there was a similar finding with respect to OUD diagnostic criteria across the three CoUD subgroups. Specifically, two diagnostic-criteria latent classes were predicted to include >70% of the individuals (split almost evenly) with one having >65% posterior probability for OUD criteria and the other having <5% posterior probability for OUD criteria.

With respect to the individual CoUD diagnostic criteria, the most pronounced difference between low- and high-PSA subgroups was for “Repeated substance use in situations where it is physically hazardous” (40% vs. 70%) while the least pronounced was for “Persistent desire/unsuccessful efforts to stop using” (93% vs. 94%). The percent difference between these two subgroups ranged from 17% to 9% for the other CoUD diagnostic criteria (Supplemental Table 6). Considering the overall criterion count, 59% of the individuals in the high-PSA subgroup met ten or more CoUD criteria while this was observed in only 34% of low-PSA subgroup (Supplemental Table 7). The frequency distribution of the CoUD diagnostic criteria in the intermediate-PSA subgroup was similar to that observed in the overall sample (Supplemental Tables 6 and 7).

### Psychiatric and somatic associations of CoUD subgroups

The analyses above demonstrated that the diagnosis-based subgroups of Yale-Penn participants with CoUD are linked to different PSA patterns. To understand their associations with psychopathology, suicidality, traumatic experiences, family history of substance use, medical conditions, and SES, we tested the three CoUD subgroups with respect to 2,952 Yale-Penn participants who did not meet criteria for any of the 5 SUDs of interest. After accounting for age, sex, and race-ethnicity and applying Bonferroni-correction for multiple testing (corrected p-value threshold < 7.94×10^−4^), we observed that many of the CoUD subgroups were associated with more severe adversity and comorbidities than the control group, with significant effects observed within each of the domains tested (Supplemental Table 8).

When comparing the associated effect sizes between the subgroups, no difference was observed with respect to the SES-related variables (difference-p>0.05), but all other categories included at least one trait with Bonferroni-corrected differences in the effects detected comparing the three CoUD subgroups to controls (Figure 2). The low-PSA subgroup had lower odds to report family/household use of any substance (OR=2.18, 95%CI=1.87-2.53) compared with either the intermediate-PSA subgroup (OR=3.76, 95%CI=3.33-4.23; difference-p=2.64×10^−8^) or the high-PSA subgroup (OR=4.66, 95%CI=4.13-5.25; difference-p=1.09×10^−14^). A significant difference between the low- and high-PSA subgroups was also present for family/household cigarette smoking (OR=2.38 vs. 3.58, difference-p=8.2×10^−5^). The three CoUD subgroups were associated with increased odds of family/household use of cocaine when growing up, but there was no difference in the strength of the associations (difference-p>0.05). With respect to psychopathology, the high-PSA subgroup was associated with higher odds of antisocial personality disorder (ASPD, OR=21.96 vs. 6.03, difference-p=8.08×10^−6^), agoraphobia (OR=4.58 vs. 2.05, difference-p=7.04×10^−4^), and posttraumatic stress disorder (OR=11.54 vs. 5.86, difference-p=2.67×10^−4^) than the low-PSA subgroup. Significant-corrected differences in effect size were also observed among the three CoUD subgroups in relation to experiencing or witnessing a major traumatic event, where the effect size was largest for the high-PSA subgroup (OR=11.54, 95%CI=9.07-14.68), and progressively smaller for the intermediate-PSA subgroup (OR=8.38, 95%CI=6.60-10.65) and low-PSA subgroup (OR=5.86, 95%CI=4.46-7.71; low-PSA vs. high-PSA difference-p=8.5×10^−16^; low-PSA vs. intermediate-PSA difference-p=3.28×10^−4^; high-PSA vs. intermediate-PSA difference-p=4.41×10^−7^). Differences were also present in the strength of the associations of the three CoUD subgroups with suicide ideation (high-PSA OR=6.1vs. low-PSA OR=3.27, difference-p=5.43×10^−9^; low-PSA OR=3.27 vs. intermediate-PSA OR=4.79, difference-p=3.29×10^−4^). Considering medical history, when compared to controls, the high-PSA subgroup was associated with increased odds of being diagnosed with sexually transmitted diseases (STD; OR=5.92 vs. 3.38, difference-p=1.81×10^−5^) and taking depression medication (OR=13.49 vs. 8.02, difference-p=1.42×10^−4^) than the low-PSA subgroup.

**Figure 2.**
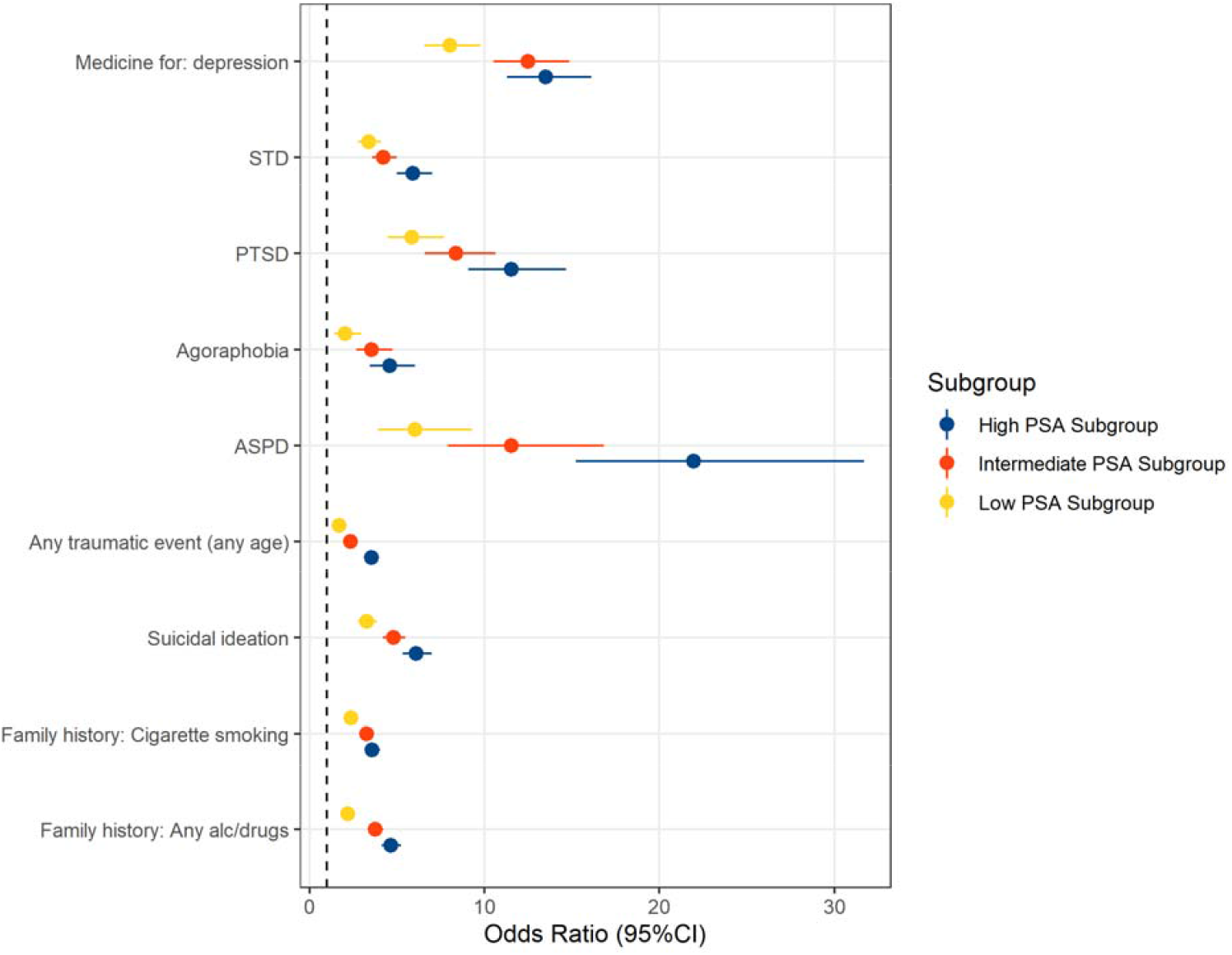
Traits with statistically significant differences across CoUD subgroups in the association strength observed when compared to the control group. Odds ratios and 95% confidence intervals (95%CI) are shown. STD: Sexually transmitted disease, PTSD: posttraumatic stress disorder. ASPD: antisocial personality disorder.

## DISCUSSION

To date, most molecular and brain imaging studies investigated CoUD-affected individuals as a singular entity, in comparison with control samples or groups of participants affected by other substance use or psychiatric disorders (50, 51). Due to the low prevalence of CoUD in the general population, few investigations of nationally representative cohorts examined differences among individuals with CoUD (52). Clinically ascertained CoUD cohorts provide a larger proportion of individuals with the disorder and offer opportunities for more in-depth clinical and phenotypic characterization. As mentioned, molecular and brain imaging studies of CoUD have mostly investigated differences between this group and controls or individuals with other psychiatric conditions (50, 51). Understanding CoUD heterogeneity in the context of PSA and commonly comorbid psychiatric and somatic comorbidities may help inform the development of more precise treatments for this population. We investigated the phenotypic diversity of 7,989 individuals with a lifetime CoUD diagnosis recruited in the Yale-Penn cohort. Leveraging this large, deeply phenotyped sample, we found that CoUD is characterized by latent typologies of individuals with different profiles of psychiatric and somatic comorbidities. Although the Yale-Penn cohort was ascertained based on the presence of addictive disorders and is not representative of the general population, our results highlight how CoUD heterogeneity could affect molecular, brain imaging, and treatment studies of addiction-ascertained samples.

Our initial LCA based on DSM-5 SUD diagnoses (i.e., AUD, CaUD, OUD, and TUD) distinguished three CoUD subgroups: low PSA (17%), intermediate PSA (38%), and high PSA (45%). In line with sex differences present among individuals with SUDs from epidemiologic studies (53), most of the individuals included in the high-PSA subgroup were males. With respect to race/ethnicity, the low-PSA subgroup has a higher percentage of non-Hispanic European Americans and high-PSA subgroup included more non-Hispanic African Americans than the overall CoUD sample. This trend parallels those reported by the National Epidemiologic Survey on Alcohol and Related Conditions-III (NESARC-III), which similarly found that non-European Americans were the majority among cocaine-only users and European Americans were the majority among cocaine-cannabis users (54). As shown in a comparison between the 2001-2002 NESARC and 2012-2013 NESARC-III, there are racial-ethnic differences among cocaine users and CoUD-affected individuals (55). Although ethnic and racial differences in CoUD are likely affected by disparities in income, education, unemployment, and discrimination (55, 56), the relationship between cocaine addiction and SES also appears to be affected by temporal trends. Indeed, although in the 1980s cocaine use was more prevalent among highly educated individuals, during the 1990s it became more prevalent among less educated individuals, and in the 2000s it increased among higher education subgroups, but not in the lowest (55). We found no major differences among CoUD subgroups with respect to income or education. This indicates that in the Yale-Penn cohort, the differences among CoUD subgroups may be influenced by cultural factors related to sex and racial-ethnic differences rather than economic differences and education.

While the general patterns observed in the LCA based on SUD diagnoses were confirmed by the LCA based on SUD diagnostic criteria (i.e., the low-PSA subgroup corresponds to CoUD with low probability of other SUD diagnostic criteria; the intermediate PSA subgroup corresponds to intermediate probability of other SUD diagnostic criteria; the high-PSA subgroup corresponds to CoUD with high probability of other SUD diagnostic criteria), we also observed some substance-specific patterns in the criterion-based LCA. For example, most participants included in the low- and intermediate-PSA subgroups were characterized by a very low posterior probability (<10%) of CaUD diagnostic criteria. Another notable substance-specific pattern was seen with OUD diagnostic criteria, where in all CoUD subgroups participants were mostly split between a class with high posterior probabilities and a class with low posterior probabilities.

Most previous studies investigating PSA in CoUD have focused on the frequency or severity of substance use (57, 58). These analyses may not have as great resolution as the analyses we performed using SUD diagnostic criteria. In the NESARC cohort, cocaine-opioid users represent about one-third of cocaine users (59). This may, at least in part, explain why a consistent number of Yale-Penn participants with low posterior probabilities of OUD diagnostic criteria were included in the CoUD subgroup with high PSA. With respect to CoUD severity, the majority of CoUD individuals with high PSA met ten or more CoUD criteria. This is in line with evidence from NESARC-III analyses where “very high quantity use” and “daily use” of cocaine were associated with polysubstance use (60). Additionally, NESARC-III analyses focused on quantity, frequency, and duration of cocaine use and identified three cocaine use patterns (61), which appear to overlap with the CoUD subgroups we observed in the Yale-Penn cohort.

In line with the well-known association of CoUD with mental and physical health (62), the three CoUD subgroups identified in our study were all associated with increased suicidal behaviors, traumatic experience, psychopathology, and somatic comorbidities relative to controls (i.e., individuals without any SUDs). However, we additionally observed several traits where the strength of the associations was statistically different among the three CoUD subgroups. With respect to family history of substance use, individuals in the low-PSA subgroup demonstrated smaller magnitude associations with family histories of alcohol and any-drug use (compared with the intermediate- and high-PSA subgroups), cigarette smoking (compared with the high-PSA subgroup), and other illegal drugs (compared with the high-PSA subgroup). Of note, the strength of the association with family use of cocaine and heroin did not differ across CoUD subgroups.

Previous studies showed that a family history of substance use is associated with polysubstance use (63-65). Our findings add to this literature and suggest that family use of multiple types of substances may be a risk factor for PSA in CoUD. Another difference among CoUD subgroups was related to suicidal behaviors. While all CoUD subgroups were associated with increased suicidal behaviors (i.e., ideation, planning, and attempt) relative to controls, suicide ideation showed smaller magnitude associations with the low-PSA subgroup, compared with the other subgroups. The association of suicidal behaviors with cocaine addiction and polysubstance use is well established (66-68). However, to our knowledge, the current study is the first to investigate suicidality in the context of PSA among individuals with CoUD. The finding that suicide ideation may be differentially impacted by PSA can be particularly noteworthy with respect to suicide prevention among high-risk individuals affected by CoUD.

Associations between CoUD, traumatic experiences, and PTSD were previously identified in multiple studies (69, 70). We found evidence for this association in the Yale-Penn cohort and observed differences in the strength of the associations across CoUD subgroups. Specifically, individuals belonging to the low-PSA subgroup were least strongly associated with having experienced a traumatic event and had the lowest odds of having a PTSD diagnosis. However, there were no differences across CoUD subgroups in their association with different types of traumatic events experiences (e.g., violence, physical abuse, and sexual abuse). With respect to other psychiatric disorders, ASPD showed the strongest associations with the high-PSA subgroup where individuals in this group had a 22-fold increase in the odds of having an ASPD diagnosis in the Yale-Penn cohort. Although low- and intermediate-PSA subgroups were also associated with ASPD, there is a strong difference in ASPD association strength between high- and low-PSA subgroups (ASPD OR= 21.96 vs. 6.03, difference-p=8.08×10^−6^). This is in line with previous studies in treatment-seeking substance users showing that CoUD and polysubstance use are both associated with ASPD with the latter having a larger effect (71, 72). Agoraphobia was the other psychiatric comorbidity showing differences across CoUD subgroups. Cocaine use and cocaine dependence have been previously associated with agoraphobia in treatment-seeking individuals and in community surveys (73, 74). We confirmed this relationship with CoUD in the Yale-Penn cohort, but also observed that CoUD with high PSA is more strongly associated with agoraphobia than CoUD with low PSA. With respect to somatic comorbidities, we observed differences among CoUD subgroups with respect to two health outcomes, STDs and the use of depression medication. There is a vast literature supporting the association of CoUD and PSA with STD risk (75-78). In Yale-Penn, the stronger STD association with the high PSA subgroup than with the low PSA subgroup is in line with these previous findings. While all CoUD subgroups were associated with traits related to depression (e.g., major depressive disorder and major depressive episodes), there was no statistically significant difference in the strength of the associations. Conversely, depression medication was more strongly associated with the high-PSA subgroup than the low-PSA subgroup. This suggests that more severe CoUD cases such as those affected by high PSA may be more frequently treated with antidepressants, despite available evidence not supporting their efficacy in treating symptoms of CoUD (79). Alternatively, the stronger association between the high-PSA subgroup and antidepressant medication treatment could reflect greater severity or chronicity of major depression or other disorders treated with antidepressants than those in the low or intermediate PSA subgroups.

Although our study contributes to the characterization of CoUD heterogeneity in a large sample, we acknowledge four main limitations. First, Yale-Penn participants were recruited for addiction genetic studies for more than 20 years. Accordingly, the associations observed may not reflect those present in the general population. As discussed above, they may be also affected by the temporal changes in cocaine use that occurred during this time span. Second, the SSADDA was designed to obtain DSM-IV diagnoses. To investigate addiction in the context of DSM-5, we derived DSM-5 SUD diagnoses from additional information collected with the SSADDA. While all DSM-5 diagnostic criteria were adequately mapped for most SUDs, two TUD criteria (i.e., social problems and neglected roles) were not available in the SSADDA. This may have affected our ability to investigate TUD comorbidity in the context of CoUD. Third, although the SSADDA is a previously validated diagnostic instrument (38, 39), the information collected may have been impacted by self-report bias (80). Fourth, the majority of the sample investigated had yearly household incomes below $40,000. This may have limited our ability to investigate the impact of socioeconomic factors on PSA of individuals affected by CoUD.

Notwithstanding these limitations, we identified three CoUD subgroups in the Yale-Penn cohort, reflecting different PSA degrees that were associated with different patterns of psychiatric and somatic comorbidities. Although the results may not generalize to general population samples, our findings nevertheless highlight the need for modeling PSA when analyzing CoUD in cohorts ascertained for addiction research such as those investigated in molecular and neuroimaging studies. Further research is needed to assess how these patterns of PSA coincide or differ in a general population. Once validated, patterns of PSA in CoUD could be assessed molecularly and with neuroimaging approaches to develop our understanding of these conditions.

## Data Availability

All data produced in the present work are contained in the manuscript and its supplemental material.

## ACKNOWLEDGMENTS

The authors thank the research participants enrolled in the Yale-Penn cohort.

## Notes

**DECLARATIONS OF COMPETING INTEREST:** RP received a research grant from Alkermes. RP and JG are paid for their editorial work on the journal *Complex Psychiatry*. JG and HRK are named as inventors on PCT patent application #15/878,640 entitled: “Genotype-guided dosing of opioid agonists,” filed January 24, 2018. HRK is a member of advisory boards for Dicerna Pharmaceuticals, Sophrosyne Pharmaceuticals, and Enthion Pharmaceuticals; a consultant to Sophrosyne Pharmaceuticals; the recipient of research funding and medication supplies for an investigator-initiated study from Alkermes; a member of the American Society of Clinical Psychopharmacology ‘s Alcohol Clinical Trials Initiative, which for the past three years was supported by Alkermes, Ethypharm, Lundbeck, Mitsubishi, Otsuka, and Pear Therapeutics, and is paid for his editorial work on the journal Alcohol: *Clinical and Experimental Research*. The other authors have no competing interests to report.

**FUNDING:** This study was supported by the National Institutes of Health (R33 DA047527, R21 DC018098, and RF1 MH132337), One Mind, and the VISN 4 Mental Illness Research, Education and Clinical Center at the Crescenz VAMC. The Yale-Penn cohort was supported by multiple grants from the National Institutes of Health (RC2 DA028909, R01 DA12690, R01 DA12849, R01 DA18432, R01 AA11330, R01 AA017535).

### Competing Interest Statement

RP received a research grant from Alkermes. RP and JG are paid for their editorial work on the journal Complex Psychiatry. JG and HRK are named as inventors on PCT patent application #15/878,640 entitled: Genotype-guided dosing of opioid agonists, filed January 24, 2018. HRK is a member of advisory boards for Dicerna Pharmaceuticals, Sophrosyne Pharmaceuticals, and Enthion Pharmaceuticals; a consultant to Sophrosyne Pharmaceuticals; the recipient of research funding and medication supplies for an investigator-initiated study from Alkermes; a member of the American Society of Clinical Psychopharmacology Alcohol Clinical Trials Initiative, which for the past three years was supported by Alkermes, Ethypharm, Lundbeck, Mitsubishi, Otsuka, and Pear Therapeutics, and is paid for his editorial work on the journal Alcohol: Clinical and Experimental Research. The other authors have no competing interests to report.

### Funding Statement

This study was supported by the National Institutes of Health (R33 DA047527, R21 DC018098, and RF1 MH132337), One Mind, and the VISN 4 Mental Illness Research, Education and Clinical Center at the Crescenz VAMC. The Yale-Penn cohort was supported by multiple grants from the National Institutes of Health (RC2 DA028909, R01 DA12690, R01 DA12849, R01 DA18432, R01 AA11330, R01 AA017535).

### Author Declarations

The studies were approved by the institutional review boards at Yale School of Medicine (APT Foundation, New Haven, CT, USA), the University of Connecticut Health Center (Farmington, CT, USA), the University of Pennsylvania Perelman School of Medicine (Philadelphia, PA, USA), Medical University of South Carolina (Charleston, SC, USA), and McLean Hospital (Belmont, MA, USA) and written informed consent was obtained from each participant.

